# Association of IL-6 -174G/C and IL-10- 1082G/A Polymorphisms and Risk of Chronicity of Hepatitis B Infection in Sudanese End Stage Renal Disease Patients

**DOI:** 10.1101/2023.01.17.23284661

**Authors:** Omer B. Mohamedsalih, Waleed A. Hussain

## Abstract

**Background:** Overt hepatitis B virus (HBV) infection is defined as infection with detectable surface antigen (HBsAg) in patient’s blood. End stage renal disease (ESRD) patients who are on maintenance hemodialysis (HD) are considered to be strong candidates for HBV infection due to prolonged vascular access via HD procedure. In addition, the differences in host immune response can be one of the reasons for the various clinical presentations of HBV infection. Polymorphisms of genes encoding the pro-inflammatory and anti-inflammatory cytokines, which are responsible for regulation of the immune response, can affect the clinical presentation of the infection. Particularly, the polymorphisms of the genes encoding cytokines such as interleukin 6 (IL6) and interleukin 10 (IL10).

**Aim:** This current study aimed to compare serum levels and allelic variant of IL-6 -174G/C and IL-10- 1082G/A polymorphisms in patients with overt hepatitis B and end stage renal disease hepatitis B patients in Khartoum State- Sudan.

**Method:** Case control study has been conducted to detect the IL-6 -174G/C and IL-10- 1082G/A polymorphisms using SSP-PCR and serum level of IL6 and IL10 using ELISA in 68 from Hepatitis B patient (37 overt hepatitis B patients and 31 end stage renal disease hepatitis B patients

**Result:** The result showed that there was no statistically difference in IL6 (174G/C) and IL10 (1082G/A) Allelic frequencies (P. value = .738, .194 respectively), Serum level (P. value = 0.36, .179 respectively) between two groups. In addition there is no significant correlation between the IL6 (174G/C) and IL10 (1082G/A) and serum level in study groups.

**Conclusion:** The Allelic Variant of IL-6 -174G/C and IL-10- 1082G/A Polymorphisms and serum levels could not play apart in complication of hepatitis B in end stage renal disease patients.

## Introduction

Hepatitis B virus (HBV) is a hepatotropic, small, enveloped DNA virus that belongs to the Hepadnaviridae family and causes an acute or chronic infection in humans. Chronic hepatitis B virus (HBV) infection represents a major global health problem, affecting an estimated 257-291 million persons worldwide and is associated with substantial morbidity and mortality because of clinical complications, such as liver cirrhosis and hepatocellular carcinoma [1].

In Sudan a recent systematic review and meta-analysis including 14 studies with 5848 participants have been conducted, revealing that HBV seroprevalence rates ranged from 5.1 to 26.8% with an overall pooled prevalence of 12.1%. According to study findings, Khartoum State had the highest prevalence of HBV infection in Sudan with a proportion of 12.7% [2].

Among many groups susceptible to HBV infection, patients with end-stage renal disease (ESRD) who are on maintenance haemodialysis (HD) are considered to be strong candidates for HBV infection due to prolonged vascular access via HD procedure at HD units where the nosocomial transmission of this virus is well documented (Ayatollahi, 2016) [3]. In addition, the impaired immune response of HD patients puts this population under serious risk of Hepatitis B [4].

Unlike the appearance of this infection in immune-competent individuals, HBV infection usually tends to be chronic in HD patients due to the immunosuppressive nature of ESRD [5]. Chronic HBV infection has three distinct states of chronicity, which can be differentiated serologically. The first one is chronic Hepatitis B characterized by detectable HBsAg in the serum for six months or more known as overt Hepatitis B. The second is an inactive HBV carrier in which the HBsAg is detectable in the serum with negative HBeAg. The third one is an unusual clinical entity known as occult Hepatitis B infection (OBI) [6].

The challenges in the area of HBV-associated disease like ESRD are the dearth of knowledge in predicting outcome and progression of HBV infection and need to understand them molecular, cellular, immunological, and genetic basis of assorted disease manifestations related to HBV infection. The differences in host immune response can be one of the reasons for the various clinical presentations of HBV infection. Polymorphisms of genes encoding the pro-inflammatory and anti-inflammatory cytokines, which are responsible for regulation of the immune response, can affect the clinical presentation of the infection. Particularly, the polymorphisms of the genes encoding cytokines such as interleukin IL-6 and IL-10 [7].

## Material and methods

### Study design and Sample collection

A case control study was conducted at Khartoum state, Sudan. Blood specimen from chronic hepatitis B patient (37 overt hepatitis B patients as control group and 31 end stage renal disease hepatitis B patients as case group) was collected in EDTA Containers and store at −20°c until processing.

### Laboratory work

#### DNA extraction

The DNA was extracted from 300 μl of blood sample using (G-spin™ Total DNA Extraction Mini Kit, intron biotechnology – Korea) according to manufacture instruction.

### Determination of DNA quality and purity

Part of the DNA solution mixed with loading dye 5 in 1 and DNA quality and purity determined using gel electrophoresis. DNA transferred into 1 ml Eppendorf tube [8].

#### DNA storage

DNA was transferred into 1ml Eppendorf tube and preserved at −20°C until PCR process.

### Polymerase chain

#### Interleukin-6 (−174G/C) and Interleukin-10(−1082 G/A) genotyping

The SSP-PCR (sequence-specific primer-polymerase chain reaction) method applied for genotyping; PCR mixture of 20 μl prepared from master mix tubes (5X FIREPOL^®^, Solis bioDyne – Estonia) for each sample. Genomic DNA amplified in two different PCRs for each polymorphism; each reaction employed ageneric antisense primer and one of the two allele-specific sense primers (see table 3.1, 3.2). To assess the success of PCR amplification in both reactions. The PCR reaction carried out in a Thermal Cycler (Techne, UK), with the following programs: 1min at 95C followed by 35 cycles of 30 sec at 95°C, 30 sec at 58°C, 30 sec at 72°C, with 5min at 72°C as final extension [9]. for Interleukin-10 genotyping, 1min at 95C followed by 35 cycles of 30 sec at 95°C, 30 sec at 55.5°C, 30 sec at 72°C, with 5min at 72°C as final extension for Interleukin-6 genotyping [8].

**Table (1).**
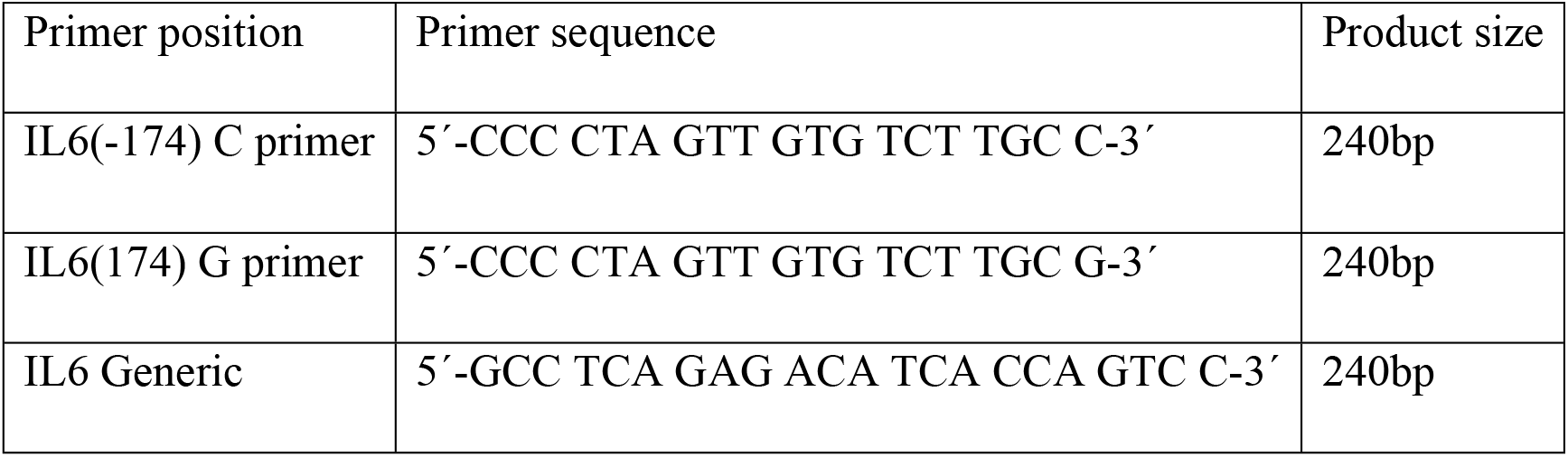
Primer Sequences Used for the IL-6 SSP Genotyping Method.

**Table (2).**
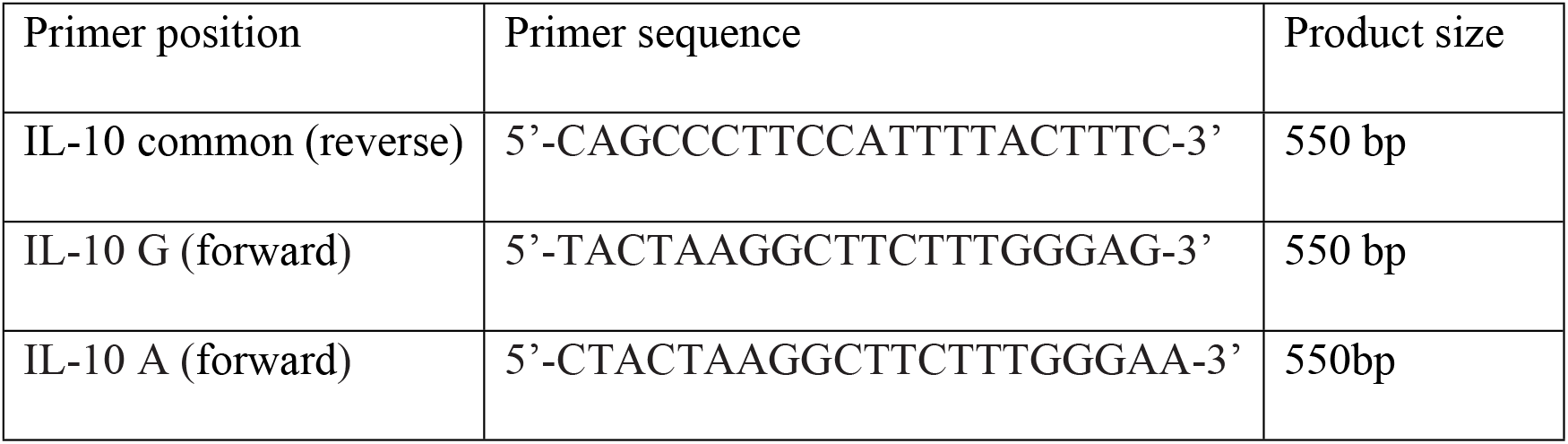
Primer Sequences Used for the IL-10 SSP Genotyping Method.

### Demonstration of PCR product

Five μl of the PCR product (ready to load) was electrophoresed on 1.5% agarose gel, and stained with ethidium bromide, 1X TBE buffer will be used as a running buffer. The Voltage applied to the gel will be 100 volt with time duration of 30 minutes. 100 pb DNA ladder will be used as molecular weight marker with each patch of samples Finally, PCR product will be demonstrated by gel documentation system [8].

### IL10 and IL6 serum level estimation

Patients serum were tested for the presence of IL-6 and IL-10 using commercially available ELISA sets for human IL-10 (ELISA MAX Standard set, #430601, BioLegend), IL-6 (ELISA MAX Standard set, #430501, BioLegend), following the protocols supplied by the manufacturers.

### Data analysis

The data analyzed using the SPSS computer program version 21. Independent sample – T. Test was used to determine the difference in genotype frequencies among study groups, the mean and stander deviation was applied for Serum level and ANOVA test with mean to correlate the genotype frequencies and serum level. The data presented in tables and figures.

## Result

In the present study we compare Serum levels and Allelic Variant of IL-6 -174G/C and IL-10- 1082G/A Polymorphisms in 31 Patients with Overt Hepatitis B and 37 End Stage Renal Disease Hepatitis B patients. The study was carried out in Khartoum state in Sudan.

Table 4.1 show that there was no statistically difference in IL6 (174G/C) Genotypes (CC, CG and GG) frequencies between overt Hepatitis B patients and end stage renal disease (ESRD) hepatitis B patients (P. value = 0.738).

**Table 4.1.**
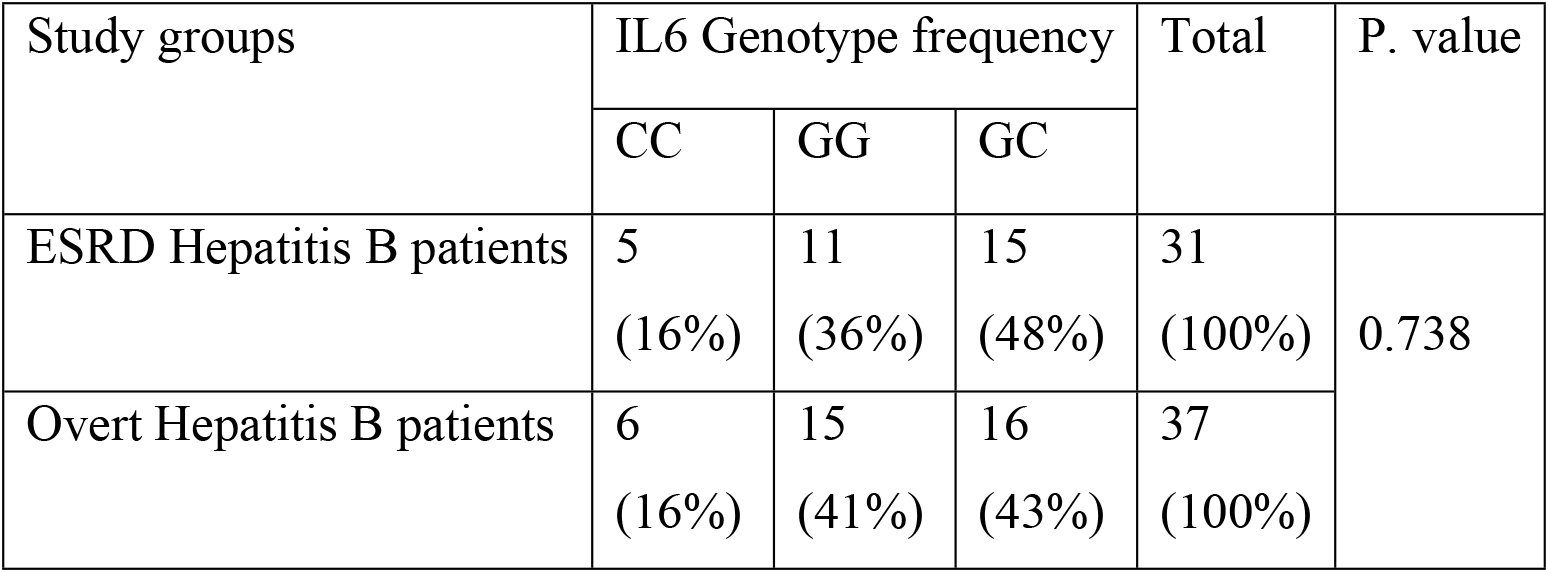
IL6 (174G/C) Genotypes frequencies among study groups.

| Study groups | IL6 Genotype frequency |  |  | Total | P. value |
| --- | --- | --- | --- | --- | --- |
|  | CC | GG | GC |  |  |
| ESRD Hepatitis B patients | 5<br>(16%) | 11<br>(36%) | 15<br>(48%) | 31<br>(100%) | 0.738 |
| Overt Hepatitis B patients | 6<br>(16%) | 15<br>(41%) | 16<br>(43%) | 37<br>(100%) |  |

Table 4.2 show that there was no statistically difference in IL10 (1082G/A) Genotypes (AA, GG and GA) frequencies between overt Hepatitis B patients and end stage renal disease hepatitis B patients (P. value = 0.194).

**Table 4.2.**
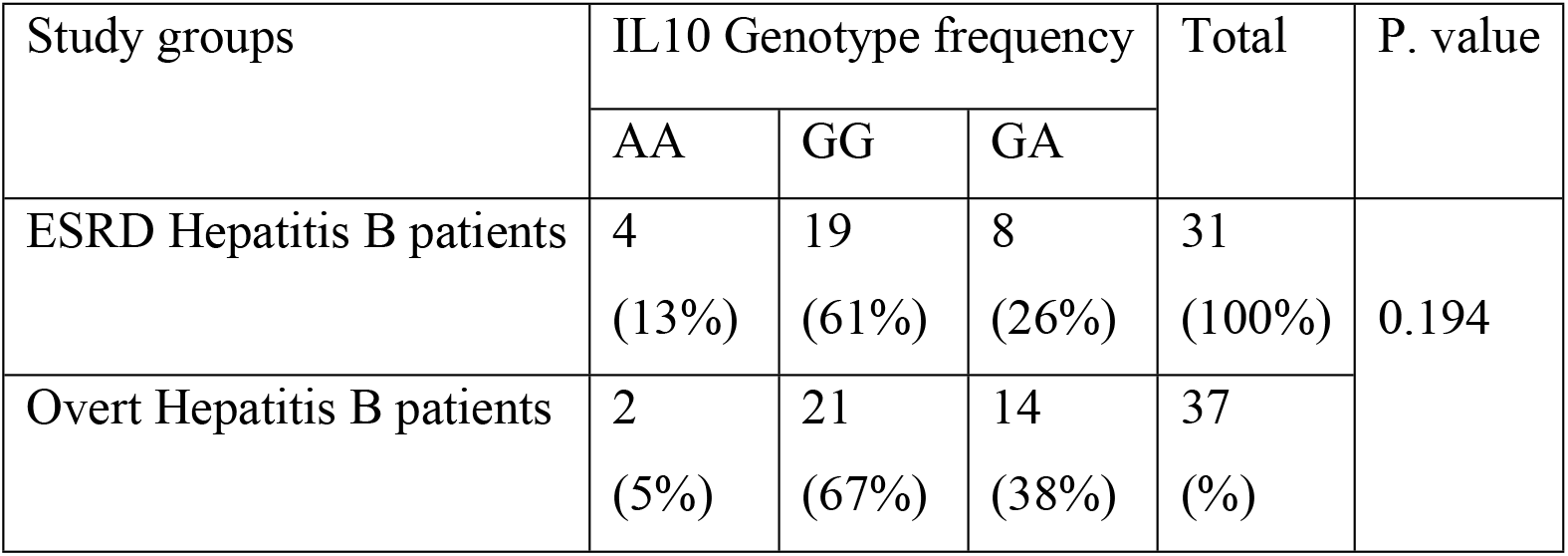
IL10 (1082G/A) Genotypes frequencies among study groups.

Table 4.3 show that there was no statistically difference in IL6 serum levels between overt Hepatitis B patients and end stage renal disease hepatitis B patients (P. value = 0.36)

**Table 4.3.** IL6 Serum Level among study groups.

| Study groups | IL6 Serum Level<br>(Mean ± STD) | P. value |
| --- | --- | --- |
| ESRD Hepatitis B patients | 15.390 ± 18 | 0.36 |
| Overt Hepatitis B patients | 9.325 ± 10 | |

Table 4.4 show that there was no statistically difference in IL10 serum levels between overt Hepatitis B patients and end stage renal disease hepatitis B patients (P. value = 0.179).

**Table 4.4.**
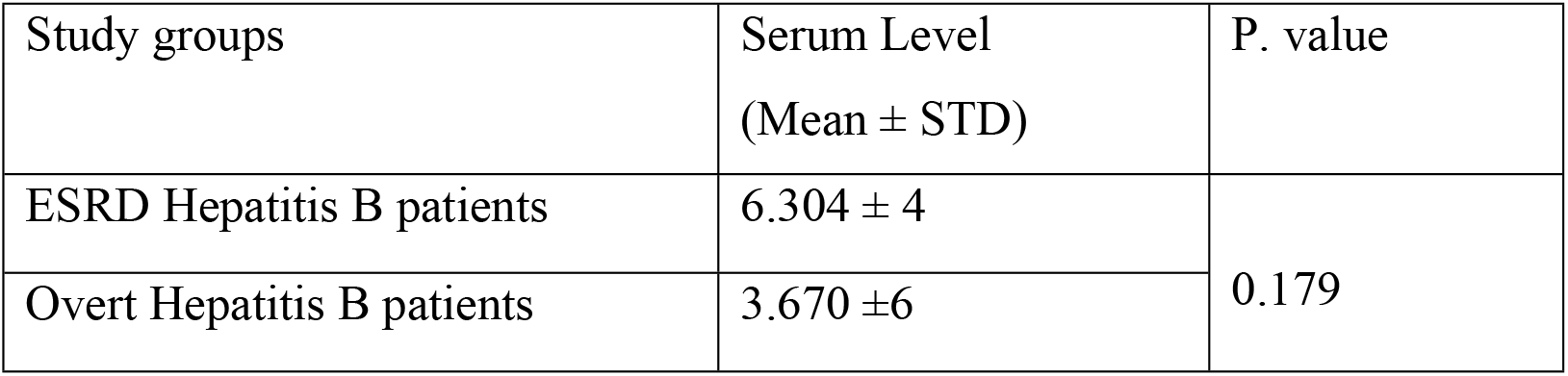
IL10 Serum Level among study groups.

Table 4.5 show that there was no statistically significant correlation between IL6 (174G/C) and serum level in Overt Hepatitis B patients and end stage renal disease hepatitis B patients (P. value = 0.536, 0.449 respectively).

**Table 4.5.** Correlation between IL6 (174G/C) polymorphisms and serum level in study groups.

| Study groups | IL6 Serum Level<br>(Mean ± STD) | | | P. value |
| --- | --- | --- | --- | --- |
|  | CC | GG | GC |  |
| ESRD Hepatitis B patients | 5.2 ± 5 | 11.7±14 | 8.5±8.2 | .449 |
| Overt Hepatitis B patients | 6.8± 9 | 18± 20 | 16±19 | .536 |

Table 4.6 show that there was no statistically significant correlation between IL10 (1082G/A) and serum level in Overt Hepatitis B patients and Overt Hepatitis B patients (P. value = 0.092, 0.229 respectively).

**Table 4.5.**
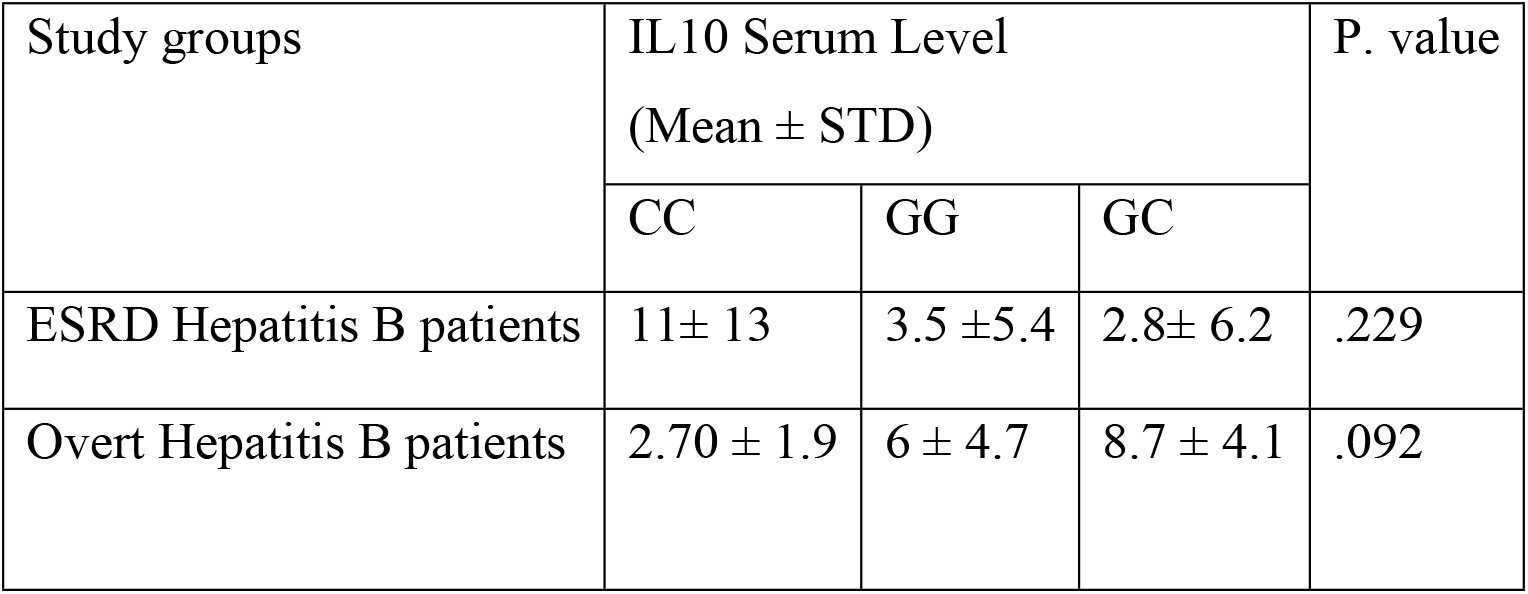
Correlation between IL10 (1082G/A) polymorphisms and serum level in study groups.

| Study groups | IL10 Serum Level<br>(Mean ± STD) | | | P. value |
| --- | --- | --- | --- | --- |
|  | CC | GG | GC |  |
| ESRD Hepatitis B patients | 11± 13 | 3.5 ±5.4 | 2.8± 6.2 | .229 |
| Overt Hepatitis B patients | 2.70 ± 1.9 | 6 ± 4.7 | 8.7 ± 4.1 | .092 |

**Figure 4.1:**
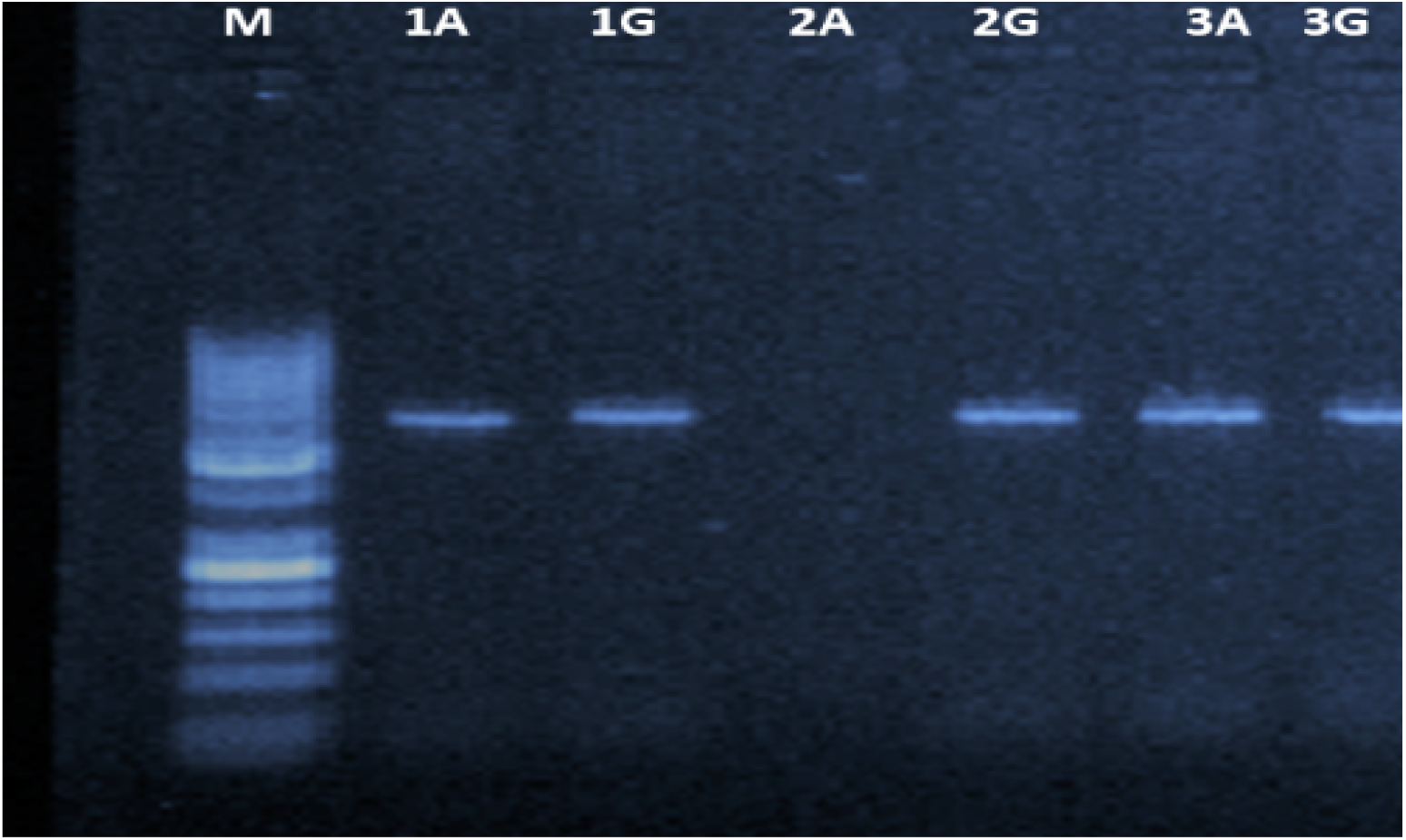
A representative Agarose (2%) gel electrophoresis of SS-PCR product (550bp) for genotyping of Interleukin-10(−1082 G/A); M: 50bp DNA marker, Lane 1, 3: represent Heterozygous mutant individuals (GA), Lane 2: represent Homozygous mutant individual (AA).

**Figure 4.2:**
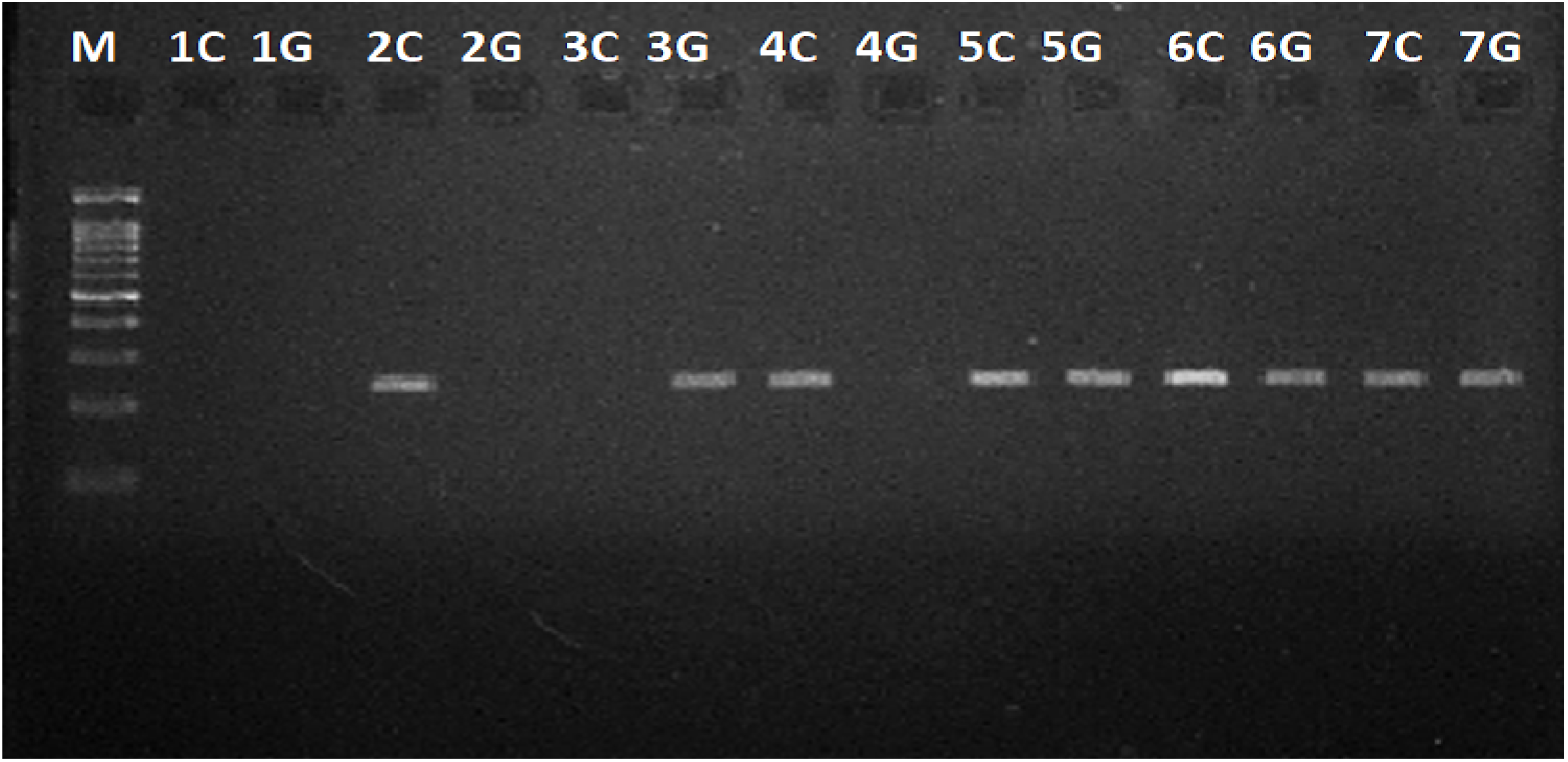
A representative Agarose (2%) gel electrophoresis of SS-PCR product (200bp) for genotyping of Interleukin-6 (−174G/C); M: 100bp DNA marker, lane 1: represent invalid sample, Lane 2, 4: represent Homozygous mutant individuals (CC), lane3: represent Homozygous wild individual (GG) and lane5, 6, 7: represent Heterozygous mutant (GC).

## Discussion

Hepatitis B virus (HBV) infection commonly induces immune reactive inflammation, which results in continuous liver tissue damage and progression of liver fibrosis to cirrhosis or hepatocellular carcinoma especially in immunocompromised patients like End stage renal disease patients [10].

Cytokines and particularly IL6 and IL10 are key regulatory elements of innate immunity against HBV infection and can affect the stage of disease. Immune dysfunction and the impaired hepatitis B vaccination response are complications of chronic renal failure that are tightly associated with inflammation induced by uremia and blood-membrane contacts [11].

Interleukin IL-6 are counter regulated by IL-10 with a large inter-individual variability. Part of the variability of cytokine production is genetically determined since polymorphisms in the cytokine gene promoters lead to high or low production [12].

The aim of this study was to compare Allelic variant of IL-6 -174G/C and IL-10- 1082G/A Polymorphisms and their serum levels in patients with overt hepatitis B and end stage renal Disease hepatitis B patients in Khartoum State-Sudan, the result illustrated that the Allelic Variant of IL-6 -174G/C and IL-10- 1082G/A Polymorphisms could not play apart in complication of chronic Hepatitis B in End stage renal disease patients, this finding is similar to meta-analysis by Ye Feng et al who conclude that the IL-6 -174G/C polymorphism has no significant correlation with the susceptibility risk of ESRD, and may not be a risk factor for ESRD [13].

Also Buckham et al. and Kandil et al. suggest that, there is no correlation between IL-6 gene polymorphism and ESRD [14, 15].

In return regarding the role of IL-6 -174G/C polymorphism in Chronic hepatitis B (CHB) and HBV related liver disease the meta-analysis done by Chang Lei et al illuminated that the IL-6 -174G/C polymorphism did not play part in susceptibility to CHB [16].

In regard to the influence of IL-10- 1082G/A Polymorphisms on immune function of CHB the study by Zhang et al showed no significant difference between IL-10- 1082G/A Polymorphisms and patient with CHB [17]. In contrast many experimental and meta-analysis studies suggest that there is association between IL-6 -174G/C and IL-10- 1082G/A Polymorphisms and their serum level and immune function of CHB patient with ESRD but all these finding awaits more verification because we do not know how the intronic variant affect the expression and level of theses cytokines.

We believe that our study has some limitation. Firstly, the sample size is relatively small, the race subgroups are missed and the collected data are not comprehensive. Finally, the influence of these promoters polymorphism on gene expression was not directly analyzed.

## Conclusion

We conclude that the allelic variant of IL-6 -174G/C and IL-10- 1082G/A polymorphisms and serum levels could not play apart in chronicity of hepatitis B in end stage renal disease patients.

## Data Availability

All relevant data are within the manuscript and its Supporting Information files.

## Recommendations

1. Direct evaluation of gene expression for these promoters polymorphism.
2. Conduct more studies regarding the intronic variant effect on gene expression and level of theses cytokines.

## Acknowledgment

Thanks to Allah almighty who enable me to stand and attend this moments, our thanks extend to all concerned persons who co-operated with me in this regard.

## Ethical consideration

This study was approved by the Ethical Committee of Scientific Research Deanship Alneelain University, and informed consent obtained from each participant before sample collection.

